# Time-Series Deep Learning and Conformal Prediction for Improved Sepsis Diagnosis in Non-ICU Hospitalized Patients

**DOI:** 10.1101/2024.11.21.24317716

**Authors:** Shaunak Dalal, Ahad Khaleghi Ardabili, Anthony S. Bonavia

**Author notes:** Corresponding author *Email address:* (Anthony S. Bonavia).

## Abstract

**Purpose:** Sepsis, a life-threatening condition from an uncontrolled immune response to infection, is a leading cause of in-hospital mortality. Early detection is crucial, yet traditional diagnostic methods, like SIRS and SOFA, often fail to identify sepsis in non-ICU settings where monitoring is less frequent. Recent machine learning (ML) models offer new possibilities but lack generalizability and suffer from high false alarm rates.

**Methods:** We developed a deep learning (DL) model tailored for non-ICU environments, using MIMIC-IV data with a conformal prediction framework to handle uncertainty. The model was trained on 83,813 patients and validated with the eICU-CRD dataset to test performance across hospital settings.

**Results:** Our model predicted sepsis at 24, 12, and 6 h before onset, achieving AUROCs of 0.96, 0.98, and 0.99, respectively. The conformal approach reduced false positives and improved specificity. External validation confirmed similar performance, with a 57% reduction in false alarms at the 6 h window, supporting practical use in low-monitoring environments.

**Conclusions:** This DL-based model enables accurate, early sepsis prediction with minimal data, addressing key clinical challenges and potentially improving resource allocation in hospital settings by reducing unnecessary ICU admissions and enhancing timely interventions.

## 1. Introduction

Sepsis is a life-threatening condition resulting from a dysregulated immune response to infection, which can lead to severe multi-organ dysfunction and death if not promptly recognized and treated [1]. Sepsis affects over 48 million people worldwide and accounts for 11 million deaths annually, making it one of the leading causes of in-hospital mortality [2]. Septic shock, a severe manifestation of sepsis, carries a mortality risk of approximately 40%, often necessitating immediate intensive care unit (ICU) admission for close monitoring and timely intervention with antibiotics, fluids, and other supportive therapies. However, patients with early-stage sepsis who are often managed in non-ICU settings are at increased risk of deterioration due to less frequent monitoring and delayed intervention. Early recognition in these patients is challenging due to the complex and variable clinical presentation of the host’s response to infection. Therefore, effective tools to identify and prioritize patients at risk of rapid deterioration and death are crucial.

Traditional methods for diagnosing and monitoring sepsis have relied on scoring systems such as the Systemic Inflammatory Response Syndrome (SIRS) criteria and the Sequential Organ Failure Assessment (SOFA) score. While these tools provide a foundation for assessing sepsis risk, they are often inefficient in detecting early stages of the disease. Recent advances in machine learning (ML), specifically deep learning (DL), offer promising alternatives by leveraging electronic health records (EHR) to predict sepsis earlier and more accurately.

The InSight ML-model has demonstrated the strong potential of using EHR data for early sepsis detection. It uses gradient boosting to predict sepsis six hours in advance of its occurrence, achieving an area under the receiver operating characteristic curve (AUROC) of greater than 0.72, and outperforming many traditional scoring systems [3]. More recent models have incorporated DL approaches, such as recurrent neural networks (RNNs) and gated recurrent units (GRUs), which leverage the temporal nature of clinical data. These models, such as DeepAISE, have shown improved predictive performance with AUROCs ranging between 0.87 and 0.90 for predictions made 4 to 12 hours before sepsis onset [4]. Transformer models have also enhanced predictive accuracy using time-series data from EHRs, achieving high accuracy while emphasizing model interpretability and transparency [5].

Despite these advances, current models for sepsis prediction face significant challenges. Many are designed for ICU settings and may not generalize well to patients in other hospital areas where data is less frequently collected [6, 7, 8]. Additionally, most models require data imputation to handle missing EHR data, which can introduce bias and reduce model reliability. Techniques such as mean imputation, forward fill, and forward/backward fill are common but do not capture the full complexity of patient data [9, 10, 5]. AI-based models also struggle with binary decision-making, leading to high false alarm rates (FAR). To mitigate the latter issue, a conformal prediction framework, such as that used by the COMPOSER model, introduces an “I don’t know” option for cases generating uncertain predictions. This approach may enhance clinician confidence in AI-driven tools by avoiding forced, potentially inaccurate decisions. [11, 12].

Considering these factors, along with the clinical need for a risk-stratification tool specifically designed for high-risk patients in a low-monitoring setting, we hypothesized that a DL-driven model that integrated time-series analyses with a conformal prediction framework could effectively address major challenges in sepsis prediction. Besides potentially improving clinician trust in AI, this approach may alleviate clinician workload while optimizing the use of critical hospital resources, such as limited number of available ICU beds, by preventing unnecessary admissions.

## 2. Results

### 2.1. Discovery and Internal Validation Using MIMIC-IV Dataset

The Medical Information Mart for Intensive Care (MIMIC-IV) database includes 328,575 non-sepsis cases and 31,207 sepsis cases, by Sepsis-3 diagnostic criteria [1]. Mean patient age was 58.8 ± 9.2 years, with 52.2% females. Patients without at least 24 hours of clinical data were excluded, resulting in 250,060 patients (8,949 sepsis and 241,111 non-sepsis).

We excluded potential model features with a high rate of missing values, to reduce model bias. This step not only streamlined model development but also ensured the model reflects the reality of incomplete data commonly encountered in the real-world setting. To that end, we assigned each laboratory type (blood chemistry panel, arterial blood gas, blood coagulation profile, and complete blood count) a missingness score of one if data were absent at the 6 h time point before sepsis diagnosis. Patients with cumulative missingness scores of three or four were excluded, resulting in a final cohort of 4,632 septic and 100,135 non-septic patients, in which the mean age for the sepsis group was 0.57 ± 0.17, 42.1% female, and the non-septic group had a mean age of 0.53 ± 0.21 with 51.0% being female. These patients were split into a training set of 83,813 patients and test set of 20,954 patients

At the 6 h, 12 h, and 24 h time windows prior to sepsis diagnosis, our Discovery model demonstrated performance parameters illustrated in Figure 1 (and Appendix Table 1), with area under the receiver-operating curve (AUROC) values of 0.99, 0.98, and 0.96, respectively (Figure 2). Although specificity for sepsis prediction remained consistently high across all time prediction windows, sensitivity decreased with an increased length of time, as expected. Given the already low false alarm rate, the conformal prediction model did not significantly reduce missed detections. More importantly, it effectively minimized false alarms.

**Figure 1:**
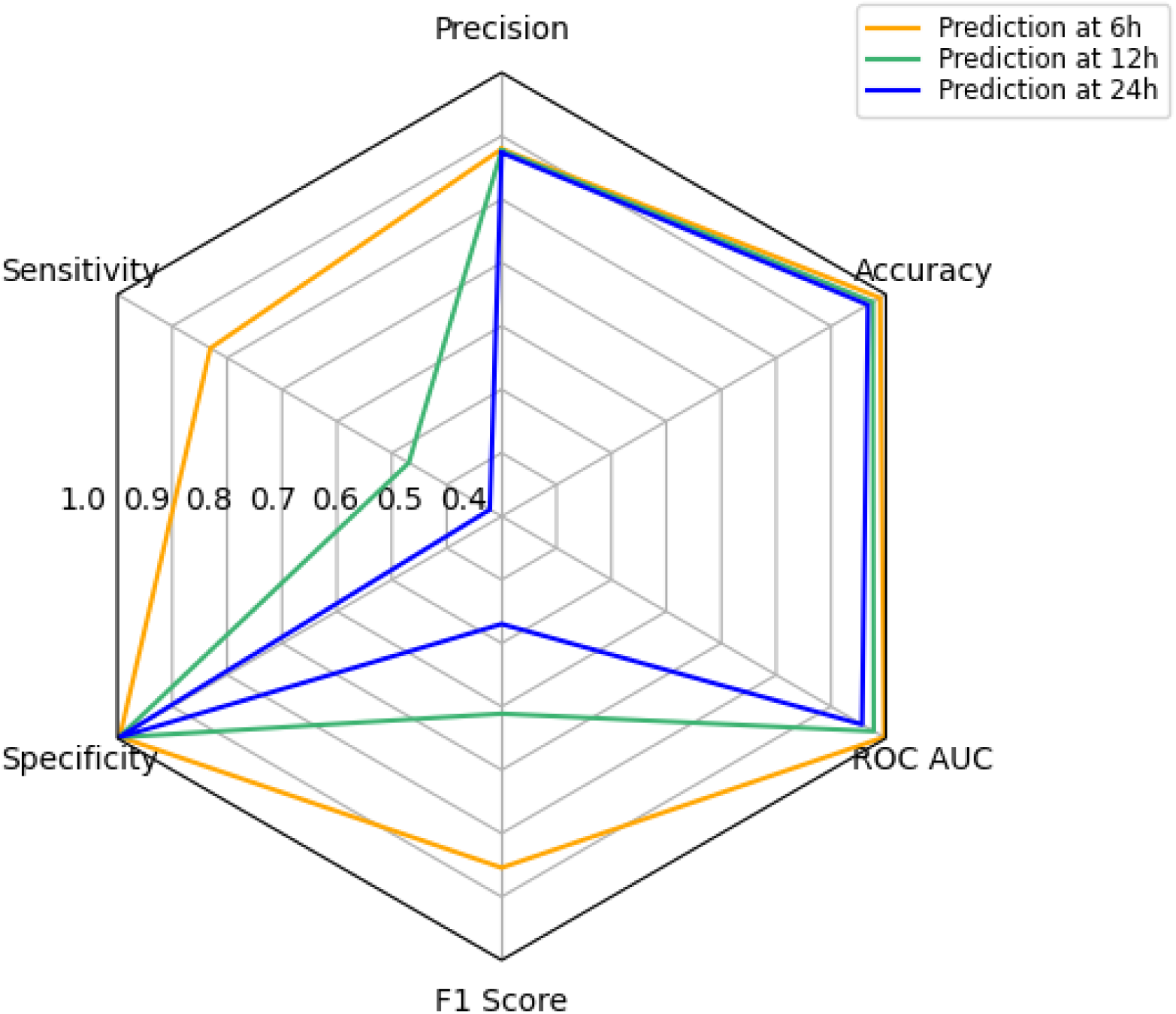
Spider Plot Comparing Model Performance at Different Time Windows using the MIMIC-IV Dataset. ‘6 h’ denotes the model incorporating data from 24 h, 12 h, and 6 h time points prior to sepsis onset. ‘12 h’ denotes the model incorporating data from 24 h and 12 h time points only.

**Figure 2:**
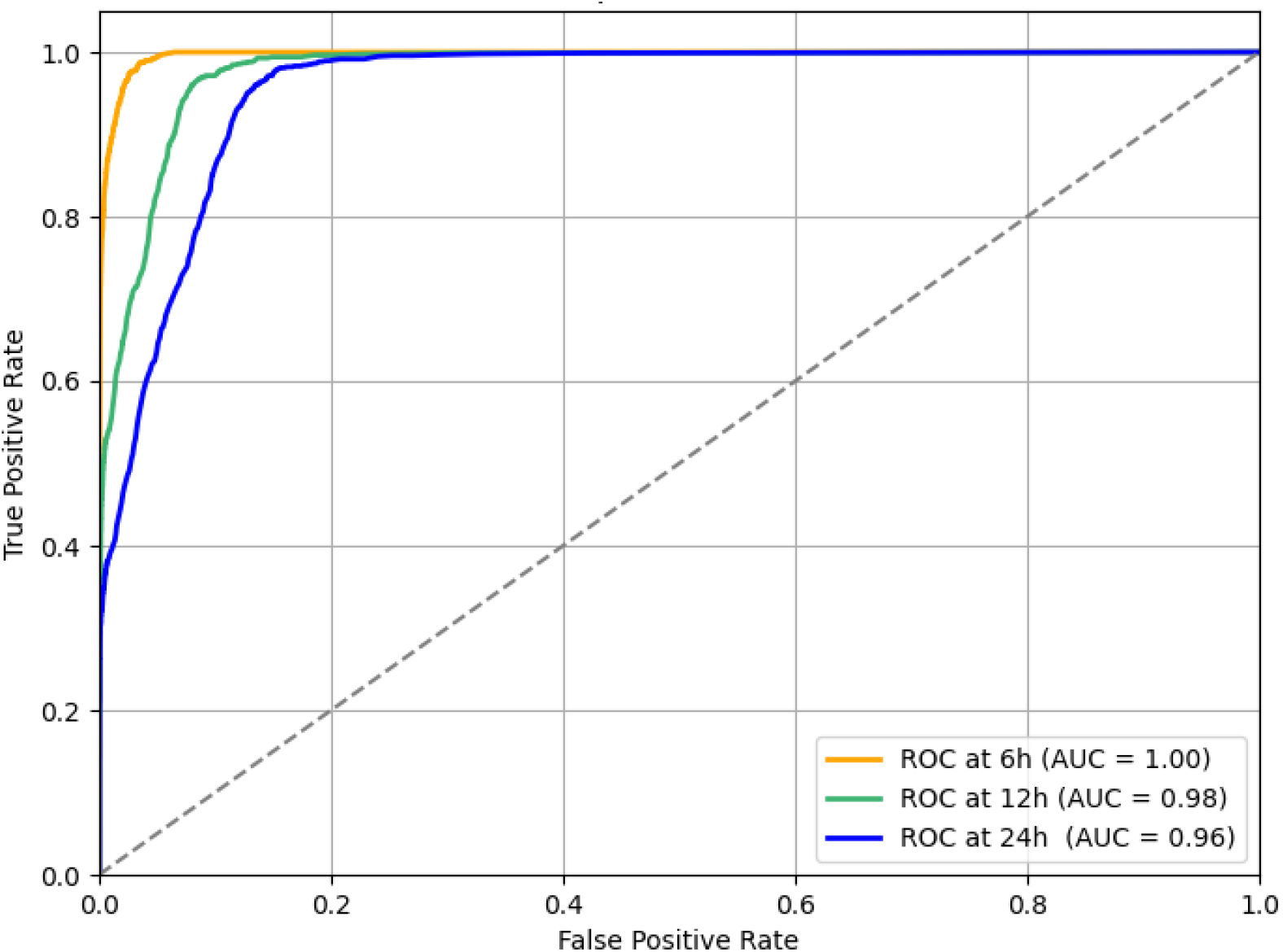
Area under the Receiver Operating Curves (AUC) at Different Sepsis Prediction Windows in MIMIC-IV Dataset. ‘6 h’ denotes model incorporating feature values from 24 h, 12 h, and 6 h time points prior to sepsis onset. ‘12 h’ denotes model incorporating features from 24 h and 12 h time points.

Models generated at time points closer to sepsis onset incorporated all available data from earlier prediction windows. For instance, at the 6 h prediction window, features with values from the 12 h and 24 h windows were included. Similarly, the 12 h model utilized features from the 24 h window if those values were available. Figure 3 illustrates that the importance of predictive features varied across the different time points. Gender, for example, played a significantly stronger role earlier in the clinical course (when fewer data points were available). However, it had minimal influence in predictions made 6 h before sepsis onset, when more important data such as leukocyte counts from 6, 12, and 24 h prior to sepsis onset were available. This dynamic performance emphasizes the importance of considering temporal evolution in feature importance when developing sepsis prediction models. As more patient data becomes available closer to the onset of sepsis, certain features that were influential earlier may diminish in significance, while others, particularly those reflecting acute physiological changes, become more critical. This dynamic adjustment highlights the need for flexible models capable of adapting to changing clinical contexts to optimize early detection and intervention.

**Figure 3:**
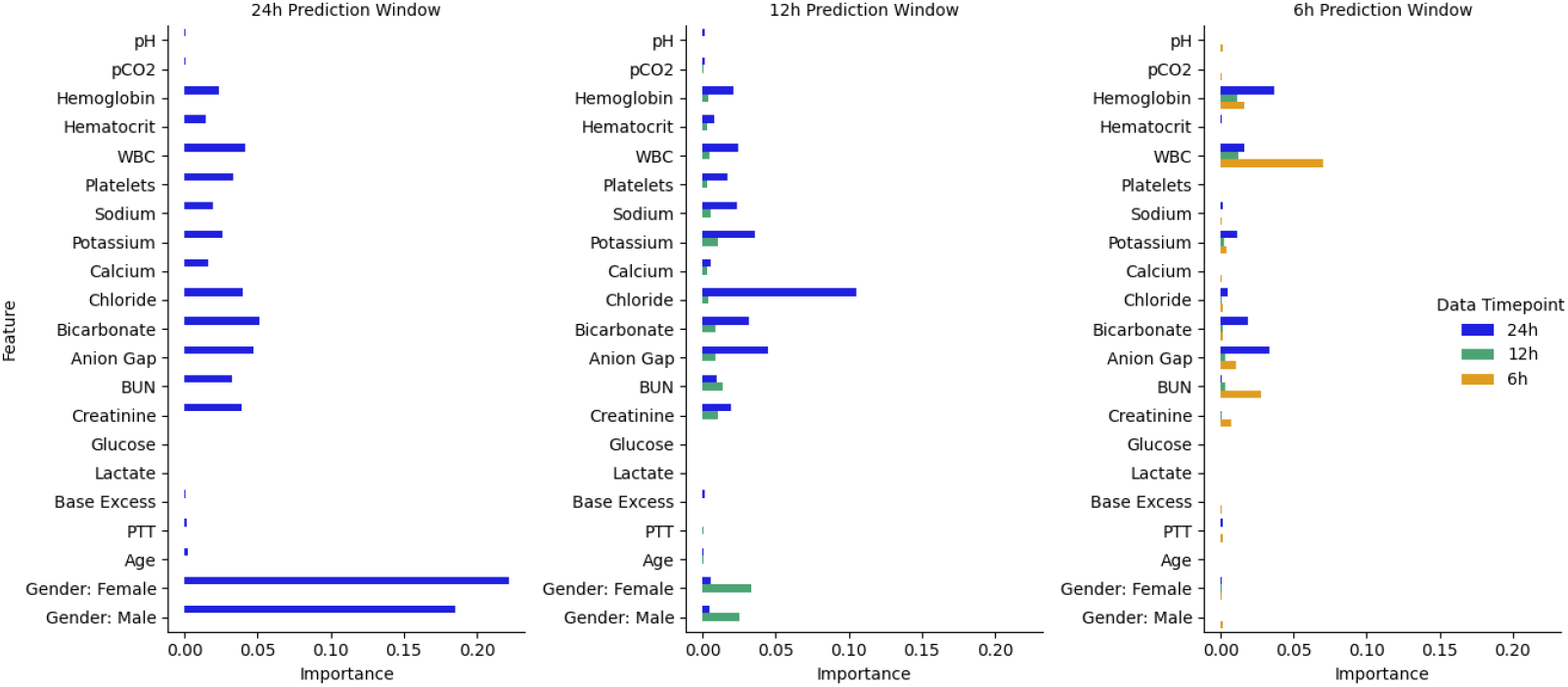
Feature importance at different prediction windows prior to sepsis onset.

### 2.2. External Validation Using eICU-CRD Dataset

Of the 195,276 patients with data in the eICU-Collaborative Research Database (CRD), 23,479 (12%) had a diagnosis of sepsis. The mean age of this Validation cohort was 64 ± 15, and 46.0% were female. Patients over the age of 89 were counted as 89 years old. Following the exclusion of patients who didn’t have clinical data from at least 24 h prior to sepsis onset, the cohort consisted of 178,137 patients (of whom 14,587 (8.1%) had sepsis). As with the MIMIC-IV dataset, we removed potential model features with high missingness, resulting in a final external validation cohort of 4,687 septic with a mean age of 0.71 ± 0.18 and 46.8% female and 77,799 non-septic patients with a mean age of 0.71 ± 0.19 and 45.2% female.

At the 6 h, 12 h, and 24 h sepsis prediction windows, AUROC values from the eICU-CRD model were 0.99, 0.98, and 0.96, respectively (Figure 4 and Appendix Table 2). However, precision and sensitivity were significantly lower when compared with performance assessed using the MIMIC-IV dataset. Notably, unlike the minimal performance enhancement observed by adding the conformal prediction framework to MIMIC-IV data, addition of this same framework caused 16% of eICU-CRD patients to have an ‘indeterminate’ sepsis prediction.

**Figure 4:**
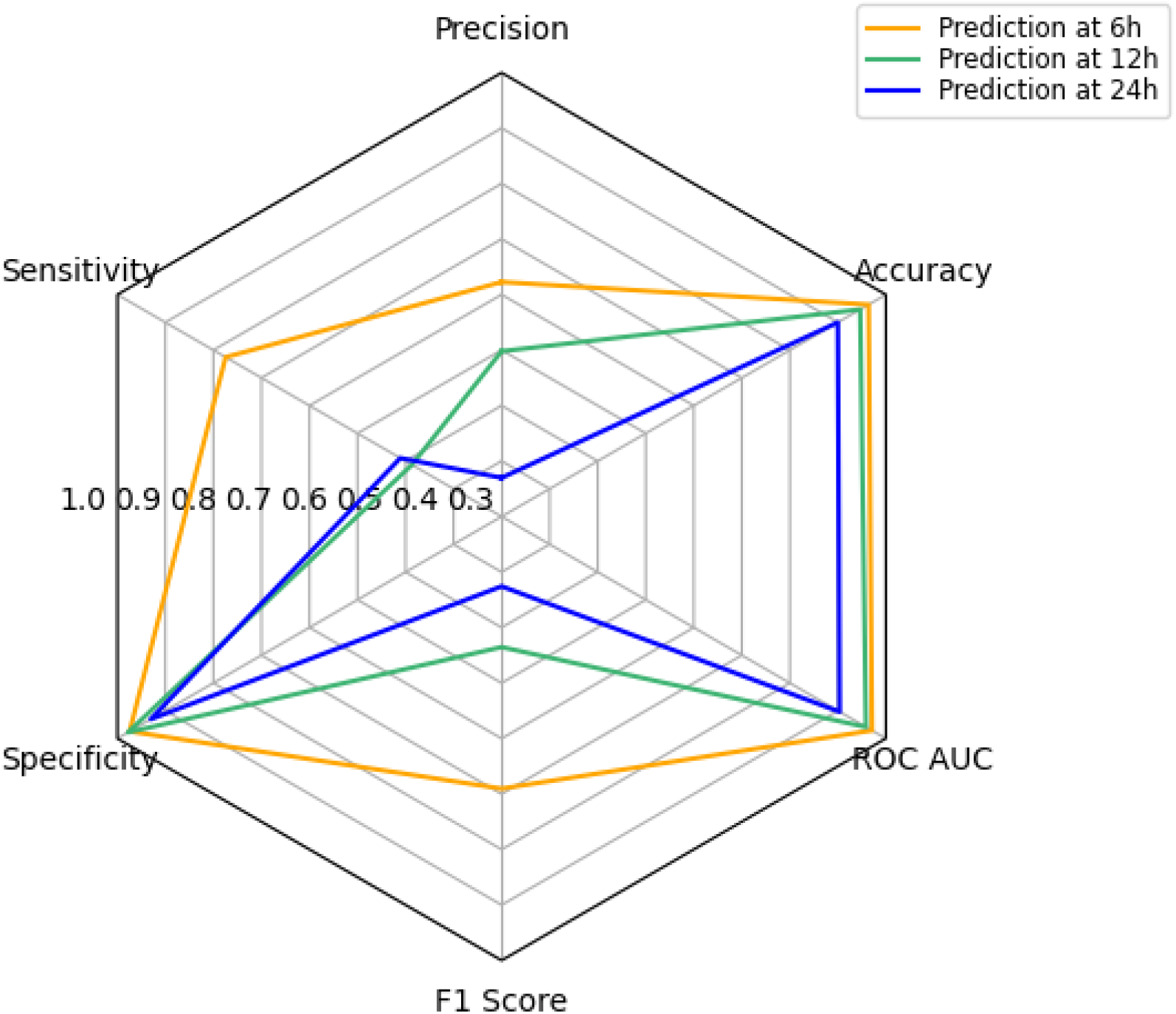
Spider Plot Comparing Model Performance at Different Time Windows in eICU-CRD Dataset. ‘6 h’ denotes model incorporating feature values from 24 h, 12 h, and 6 h time points prior to sepsis onset. ‘12 h’ denotes model incorporating features from 24 h and 12 h time points.

We also observed a 4% decrease in the FAR with the use of the conformal prediction framework as well as an increase in specificity across each time window (Figure 5). At the 6 h time point, there was a 57% reduction in false positive predictions and a 20% reduction in false negatives, and a similar pattern was observed across subsequent time points.

**Figure 5:**
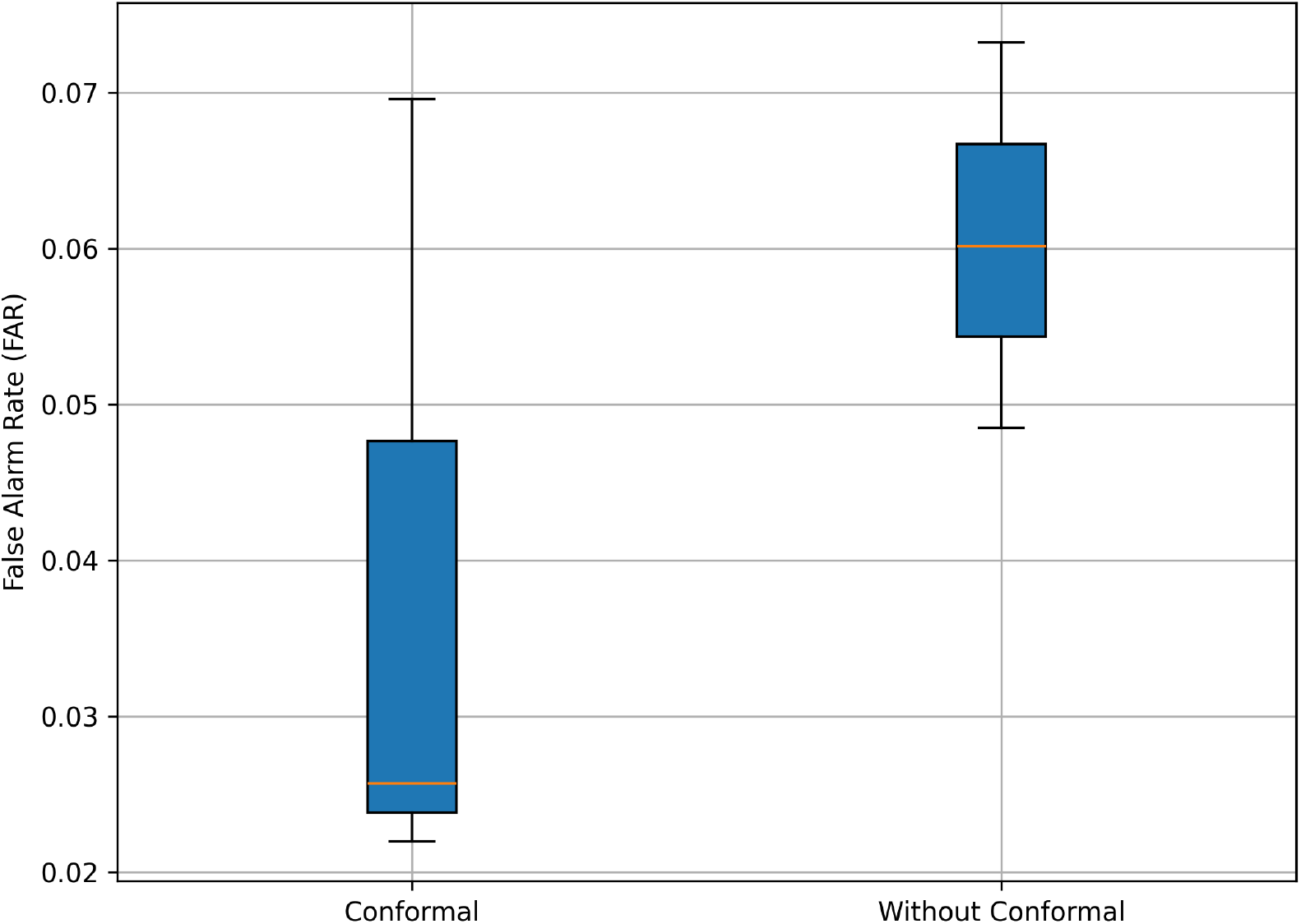
False Alarm Rate (FAR) comparison when using conformal prediction on eICU-CRD

## 3. Discussion

Despite substantial investment in sepsis healthcare and research over the past few decades, there has been limited progress in developing new disease therapies [13, 14, 15] or prognostic tools [16, 17]. The well-established principle of early detection for prompt implementation of sepsis bundles therefore remains the essential cornerstones for reducing sepsis-associated morbidity and mortality [18]. The present analysis directly addresses this continued need to identify high-risk patients for early implementation of these clinical bundles, while being cognizant of existing barriers to the widespread implementation of AI-driven clinical tools. We have demonstrated that an RNN-based risk-stratification tool that uses easily available data parameters can identify patients who will develop sepsis with a high degree of precision, even 24 hours in advance of diagnosis. This time window provides ample time for clinicians to develop and enact diagnostic and therapeutic measures to improve outcomes in these high-risk patients. We have further demonstrated that our results could be generalized to a completely different patient population, making this approach clinically feasible.

While artificial intelligence (AI) tools are beginning to be integrated into clinical practice [12], their adoption is hindered by clinicians’ concerns about false alarms, imperfect algorithms and limited transparency[19]. In direct response to these barriers, we have developed a diagnostic tool specifically optimized for use with the limited data that is available for patients receiving care outside an ICU settings—those same patients who are most vulnerable to delays in sepsis care due to undetected clinical deterioration. Our time-series model predicts sepsis at 24 hours prior to its occurrence with precision, accuracy, and specificity comparable to models based on data measured just 6 hours prior. Key features of our model include the use of a minimal set of inputs and the incorporation of a recently described conformal prediction framework that prioritizes an “I don’t know” response over false predictions [11]. We believe that these attributes are responsive to major physician concerns regarding AI-guided clinical care. Furthermore, we propose a conceptual framework that would make similar tools valuable in a patient care setting (Fig. 6).

**Figure 6:**
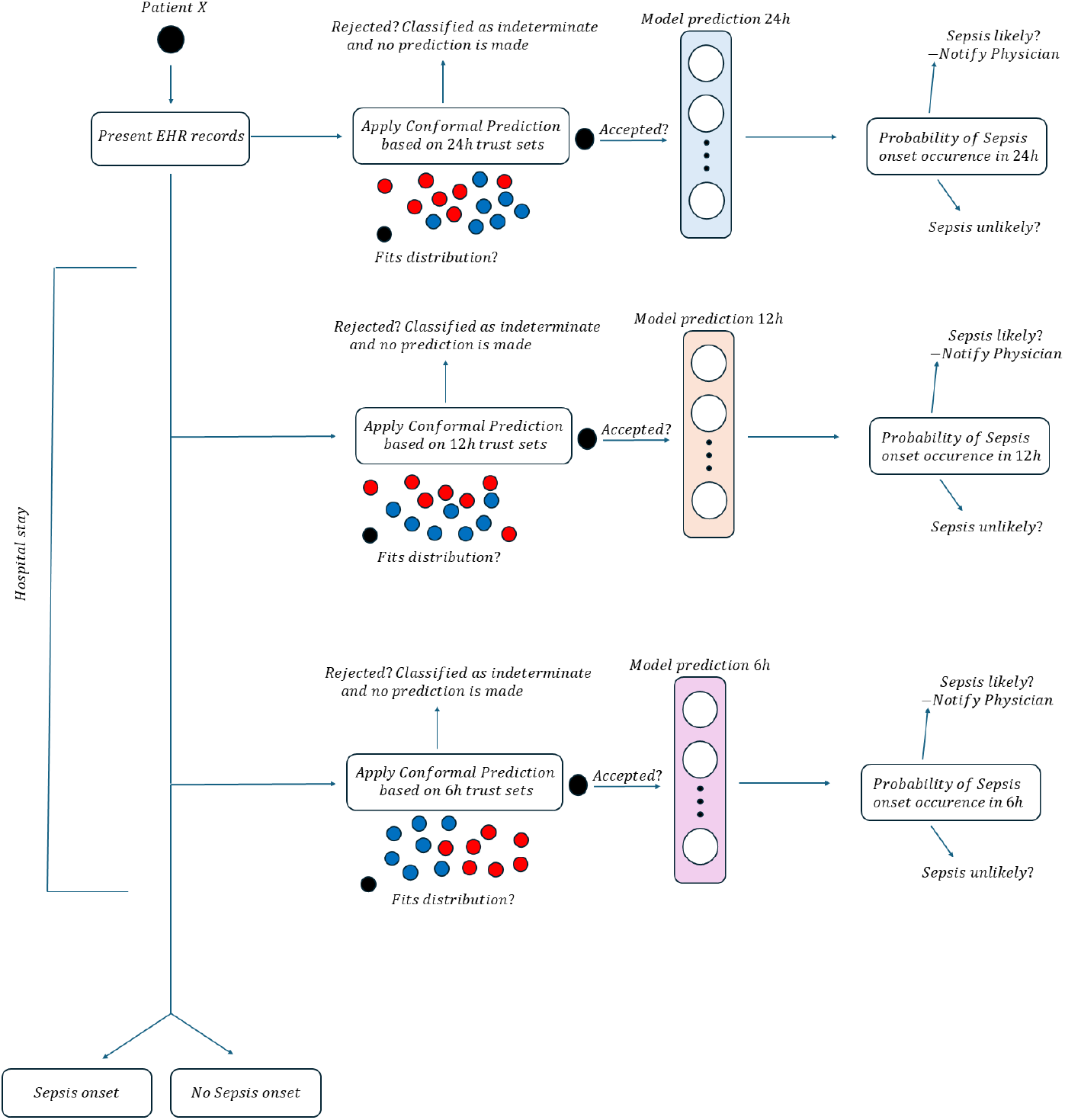
Proposed Systems-Level Use Implementation of Sepsis Prediction Model in the Setting of an Integrated Healthcare Setting.

External validation is one of the most challenging and time-consuming components of ML-based model prediction. While EHR-based ML studies have focused on avoiding pitfalls that lead to unreliable model generation (e.g., data missingness, inappropriate patient inclusion/exclusion, prediction bias), [20, 21] external model validation is still notably lacking in most studies. In the field of infectious diseases, a systematic review identified 232 prediction models for the diagnosis and prognosis of COVID-19 infection [22].

However, only two of these models were validated through collaborative efforts and data sharing [22]. Another systematic review of 34 publications that examined ML methods in clinical settings reported that only two performed external validation [23]. These findings may explain a lack of buy-in by healthcare professionals regarding the generalizability of ML predictions, and were a specific focus of the present investigation.

MIMIC-IV and eICU-CRD databases differ notably in the patient populations they describe. MIMIC-IV contains data from a single tertiary academic medical center—the Beth Israel Deaconess Medical Center in Boston, Massachusetts—collected over a period from 2008 to 2019. This data represents a more homogeneous patient population with standardized clinical practices and consistent data recording, but it is localized to an urban setting and may not capture the regional variability found elsewhere. In contrast, eICU-CRD includes data from over 200 hospitals across the United States, collected during 2014–2015 and capturing a broad and diverse patient population with variations in geography, hospital types, and clinical practices. While eICU-CRD’s nationwide scope enhances the generalizability of research findings, it introduces complexity due to variability in data recording and clinical practices compared to the consistency found in MIMIC-IV. The most relevant difference between datasets for our application, however, was the fact that MIMIC-IV allowed us to identify septic patients according to current, Sepsis-3 criteria [1], while the eICU-CRD did not. Hence, we relied on EMR-listed problems to identify sepsis patients within the latter dataset. This was a limitation of our study, although it also allowed us to leverage the conformal prediction framework to full effect.

Our conformal prediction framework was based on the approach recently described by Shashikumar et al. [11]. Trust sets were constructed by selecting cases based on levels of data missingness and cross-entropy error, focusing on those that achieved the highest F2 score. Unseen data was subsequently assessed for “typicalness” with respect to the trust set. If the *P* -value for any differences fell below a predefined threshold, the hypothesis that unseen data fits within the same distribution of the trust set is rejected, leading to an “I don’t know” response. This simple but effective approach improved the FAR by only *<*1% in the MIMIC-IV test dataset, although almost 17% of patients in the eICU-CRD were labeled as indeterminate for a diagnosis of sepsis. This translated, for example, into a 2.2% increase in specificity and a 93% decrease in the FAR at the 12 h prediction window.

Our study has several limitations, including its reliance on laboratory data. Vital signs are key physiologic indicators of illness, although they were not available in MIMIC-IV for patients receiving care outside the ICU setting. We prioritized the clinical need for identifying early-sepsis patients over the inclusion of vital signs, being cognizant of the more dire clinical repercussions of missing a diagnosis of evolving sepsis in a less monitored clinical setting.

Second, while the conformal prediction framework integrated into our model helps reduce false-positive and false-negative predictions by providing an “I don’t know” response, this approach also introduces ambiguity into clinical decision-making. The potential impact of this uncertainty on clinical workflows and decision-making processes has not been fully assessed and may require further exploration.

Third, our model’s reliance on time-series data introduces challenges related to data completeness and quality. Although we implemented a custom masking layer to handle missing data, our approach still depends on the availability of sufficient and reliable historical data to generate accurate predictions. In real-world clinical environments, data entry errors, variations in laboratory test ordering, and other data quality issues could affect the model’s performance.

## 4. Conclusion

The present analysis introduces an AI-driven model informed by EMRs to address an important clinical need. While the performance metrics demonstrate the model’s ability to reduce false alarms and missed detections, its novelty lies in its potential applicability in non-ICU settings, its effectiveness with incomplete datasets, and its reliance on temporal data. This work illustrates how the effective implementation of ML, when guided by clinical needs, can be designed to enhance patient care.

## 5. Methods

### 5.1. Data Collection

To train and internally validate our sepsis prediction model, we used the MIMIC-IV dataset v2.2 (accessed June 10, 2024). This relational database contains patient information both within and outside the ICU, for any given hospital admission at Beth Israel Deaconess Medical Center. Vital signs are only included for ICU stays, and given that our goal was to develop an effective, ‘early alert’ tool with clinical utility outside the closely monitored ICU setting, vital signs were therefore not included as features in our predictive models.

The primary outcome variable was a diagnosis of sepsis using Sepsis-3 criteria [1]. Thus, the model was trained to predict sepsis at 24 h, 12 h, and 6 h prior to diagnosis of the primary outcome. For non-septic controls, a ‘non-sepsis’ time was randomly generated such that at least 24 h of clinical data for that hospitalization was available prior to this time. For septic patients, this meant ensuring the patient had a hospital stay of at least 1 day until sepsis onset.

Laboratory data for the feature of interest within 6 h prior to the time point of interest was used for model training. For example, if serum creatinine concentration was not measured at exactly 12 hours prior to sepsis diagnosis, the algorithm searched for the temporally closest, measured value between 12 - 18 h prior to sepsis.

We generated our validation dataset by applying the same extraction criteria to the eICU-CRD and using the same model features and time windows. Due to limitations in data availability, we identified septic patients using diagnosis strings. In the eICU-CRD dataset, the diagnosis string, along with the ICD-9 Code, is a text field that categorizes and describes a patient’s diagnosis, progressing from broad categories to specific terms. For example, a patient’s diagnosis string might read: “infectious diseases—systemic/other infections—signs and symptoms of sepsis (SIRS)—due to infectious process with organ dysfunction.” Patients with the term “sepsis” in their diagnosis string were classified as septic.

### 5.2. Data Pre-processing

We removed model features having a high rate of missingness in one or more of the following categories: blood chemistry, arterial blood gas, blood coagulation parameters, or complete blood count. Every system missing data at the 6 h time point prior to sepsis diagnosis was assigned a cumulative missingness score of one. The 6 h time point was selected to account for the fact that data is most likely to be available at the time point that is most proximal to the primary outcome. Based on the distribution of these scores from all patients, we eliminated patients with missingness scores of two or three. The final feature set consisted of 18 dynamic features and two demographic variables.

Further data correction techniques included winsorization (Winsor coefficient of 0.05%) to reduce the impact of outliers and consideration of variance inflation factors and correlation scores to ensure data robustness. Features exhibiting high skewness and kurtosis, such as white blood cell counts, were log-transformed. We normalized all numeric features, including age, and used one-hot encoding to convert gender into binary columns. To support our time-series-based deep learning approach, we avoided traditional imputation. Instead, we filled missing values with -1, allowing the model to handle these independently through a masking layer that skips over the time points for a missing feature, preventing the model from learning from that pattern. We employed a train-test split of 80:20 for internal model validation.

### 5.3. Model Construction

#### 5.3.1. Masking Layer

To address persistent data missingness in the selected model features, we implemented a custom masking layer. This layer effectively ignores missing values by skipping the corresponding time point for that patient, thus ensuring that the model is not biased by incomplete training data. We designated missing values as -1 (the mask value) to enable the masking layer to easily identify them. This approach allowed us to conduct time series analysis without relying on extensive data imputation and associated model bias.

#### 5.3.2. Weighted Input Layer

Clinicians frequently order additional diagnostic tests as a patient’s condition deteriorates, focusing on more recent data to make timely decisions. To emulate this clinical practice, we designed a custom weighted input layer in our model that assigns higher weights to measurements taken closer to the sepsis prediction window. This weighting is based on “delta time” values, which quantify the time elapsed before the prediction window. For example, in a 6 h prediction window, measurements taken at 6, 12, and 24 h before sepsis onset would have delta time values of 0, 6, and 18 h, respectively. This method prioritizes more recent and relevant data, thereby improving the model’s predictive accuracy.

The layer uses two trainable parameters, *α* and *β*, to uniformly initialize the weights, where *α* controls the importance of each lab feature, and *β* is used to scale the weights. The weights are computed using an exponential decay function based on *α, β*, and the delta values. Specifically, for each feature measured at different time points, the weight is calculated as follows:

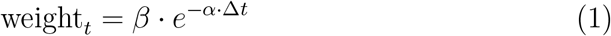

where Δ*t* represents the time delta before the sepsis prediction window and *t* represents the sepsis prediction window (24 h, 12 h, 6 h). The weighted features for each time point are concatenated with demographic, non-weighted features. This results in the final input tensor for the model.

#### 5.3.3. Recurrent Neural Network

Given the time series-based sepsis prediction task, recurrent neural networks (RNNs) are well-suited because they can effectively model sequences and handle time series data. RNNs excel in capturing both long-term and short-term dependencies, which are crucial for analyzing the complex relationships between sepsis predictors and their respective time points.

Our approach incorporated an encoder-decoder framework with a Long Short-Term Memory (LSTM) layer as the encoder with 64 units. The layer processes the input sequence step by step, retaining information through memory cells. Then, the LSTM encoder returns both the sequence of hidden states for attention processing and the final hidden and cell states, which summarize information from all previous steps to capture long-term dependencies across the entire input sequence. This enables the model to learn and retain important patterns in the data over time.

We integrated an attention mechanism to enhance the model’s focus on relevant time steps. Moreover, learned attention weights can be used to interpret the output risks, which is helpful for application to practical clinical settings. The self-attention mechanism takes the outputs of the encoder LSTM and computes a weighted sum. This mechanism allows the model to hone in on past and present features indicative of sepsis, thereby improving prediction accuracy.

The decoder consists of a fully connected dense layer of 64 units with Rectified Linear Unit (ReLU) activation, followed by a sigmoid activation function. The attention-enhanced context vector is passed through this dense layer, which refines the information and helps generate the final output. The sigmoid activation at the output layer produces a sepsis risk probability ranging from zero to one, indicating the likelihood of sepsis onset for each patient (Fig. 7). The model was compiled with the Adam optimizer, a learning rate of 0.001, and used binary cross-entropy as the loss function.

**Figure 7:**
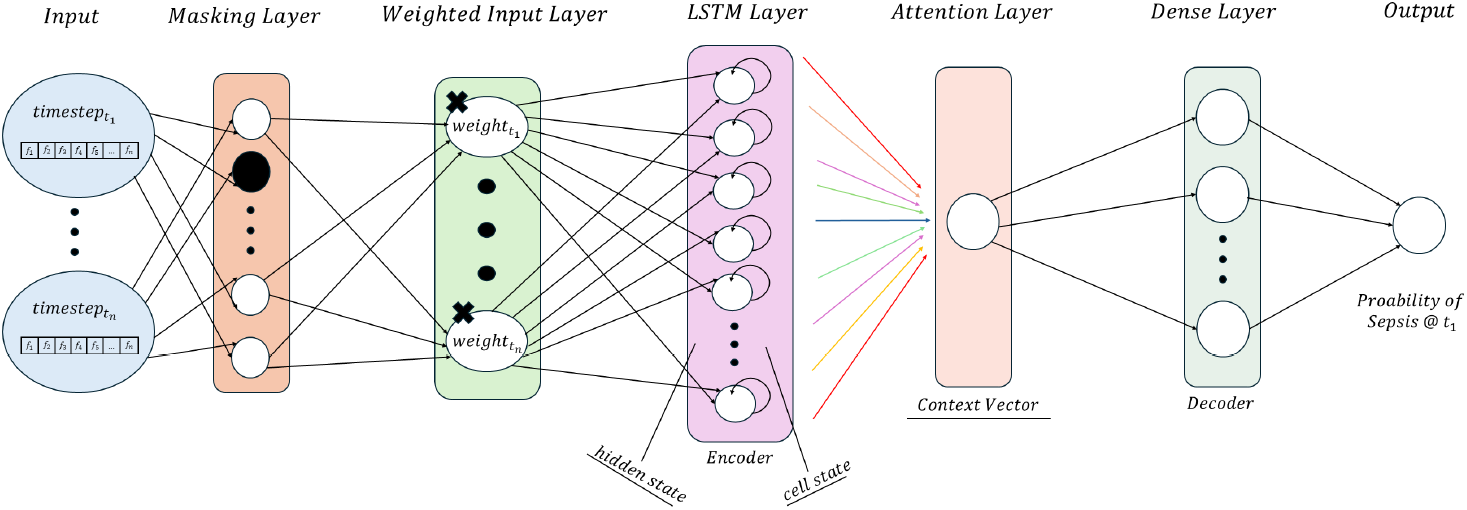
Predictive Model Architecture Using Time Series Input

### 5.4. Conformal Prediction Framework

To mitigate the effect of AI overconfidence in clinical practice, where false alarms in the hospital may increase resource consumption, overload hospital staff, and hinder clinical buy-in, we incorporated a conformal prediction framework within the context of our temporally aware sepsis prediction algorithm. This framework was based on the model employed by Shashikumar et al.’s COMPOSER (COnformal Multidimensional Prediction Of SEpsis Risk) model [11].

After training the prediction model at each time point, we focused on cases where the model predictions had maximized error in the respective sepsis and non-sepsis patients. Specifically, clustered non-sepsis patients with predicted high-risk scores and sepsis patients with low predicted scores were grouped. Thereafter, we filtered these two groups only to include patients with more than 20% missing data. This methodology created two trust sets of size *N*, randomly under-sampled. The conformal sets were used to determine how similar a newly shown patient is to the model’s training data at a specified prediction time (Fig. 8).

**Figure 8:**
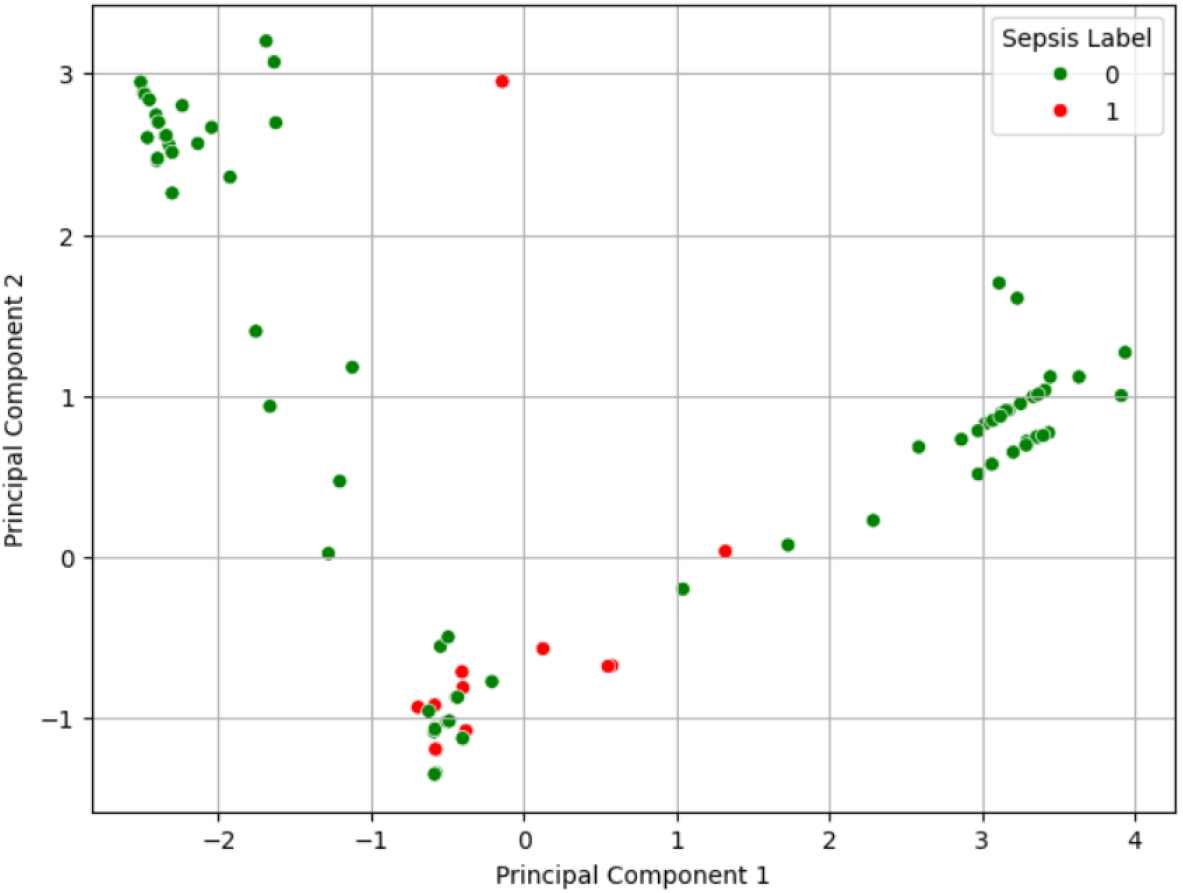
Dimensional analysis of the conformal sets at the 12 h prediction window.

We used a nonconformity measure by taking a test example and computing the average Euclidean distance between that sample and all other samples in the trust sets. The *p-*value represents the typicalness of the test sample within the probability distribution of the respective trust set. To calculate the *p-*value, the test example is added to the trust sets, and nonconformity scores for all examples in the combined set are computed. Subsequently, the proportion of nonconformity scores in the combined set that are greater than or equal to the test sample’s score is calculated.

Formally, let *τ*_septic_ and *τ*_non-septic_ denote the trust sets for septic and non-septic cases, respectively. For a test example *x*, we define the nonconformity score *η* as:

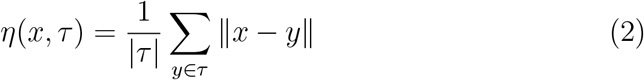

Where ∥ *x* − *y* ∥ is the Euclidean distance between *x* and *y*.

The *P-*value for the test example *x* with respect to the trust set *τ* is then calculated as:

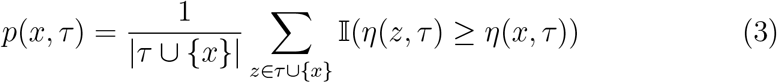

where 𝕀 is the indicator function.

For each test sample, the *P-*value is calculated with respect to both trust sets, *τ*_septic_ and *τ*_non-septic_. If the *P-*value is greater than our selected significance level (*ϵ* = 0.05) for either trust set, the example is conformant (accepted), meaning it falls within the same probability distribution of at least one of the trust sets. Mathematically, this can be expressed as:

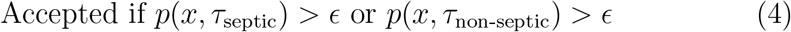

If the *P-*value with respect to both trust sets is less than *ϵ*, then the sample is non-conformant (rejected), meaning it does not fit in either septic or non-septic distribution:

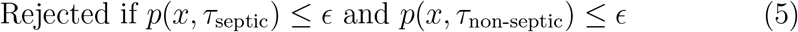

Accepted samples proceed to the sepsis prediction algorithm for a prognostic risk score, while rejected samples are classified as indeterminate.

## Data Availability

All data produced in the present study are available upon reasonable request to the authors.

## 6. Acknowledgements

We would like to thank Drs. Shamim Nemati and Supreeth Shashikumar for providing a MATLAB-based implementation of the conformal prediction module for a simulated data set.

## 7. Declarations

### Ethics approval and consent to participate

De-identified and publicly-available human data was accessed via Physionet [24] and utilized in accordance with PhysioNet Credentialed Health Data Use Agreement 1.5.0. The eICU-CRD database was similarly utilized in accordance with the corresponding Data Use Agreement [25].

### Author Contributions

A.S.B. conceived and designed the study. S.D. and A.K.A. developed the methodology and performed the data analysis. S.D. and A.K.A. contributed to data acquisition and curation, and wrote the applicable code. A.S.B. provided funding, resources, and supervised the project. S.D. and A.S.B. wrote the original draft of the manuscript, and all authors contributed to reviewing and editing the manuscript. All authors read and approved the final version of the manuscript.

### Code Availability

Code is available by reasonable request from the corresponding author.

### Data Availability

Both MIMIC-IV and eICU-CRD datasets are publicly available.

### Consent for Publication

Not applicable.

#### 8. Glossary

Sepsis: A life-threatening condition arising from the body’s dysregulated immune response to an infection, potentially leading to tissue damage, organ failure, and death.
Systemic Inflammatory Response Syndrome (SIRS): A clinical syndrome characterized by systemic inflammation.
Sequential Organ Failure Assessment (SOFA): A scoring system that assesses the extent of a patient’s organ function or rate of failure.

## Appendix A Supplementary Material

**Table A.1:** Performance Metrics for Predictions at 6h, 12h, and 24h Before Sepsis Onset in MIMIC-IV Dataset.

| Prediction Time | Precision | Sensitivity | Specificity | F1 Score | ROC AUC | Accuracy |
| --- | --- | --- | --- | --- | --- | --- |
| 6h Window | 0.879 | 0.830 | 0.995 | 0.854 | 0.997 | 0.989 |
| 12h Window | 0.877 | 0.469 | 0.997 | 0.611 | 0.978 | 0.976 |
| 24h Window | 0.874 | 0.321 | 0.998 | 0.470 | 0.956 | 0.967 |

**Table A.2:** Performance Metrics for Predictions at 6 h, 12 h, and 24 h Before Sepsis Onset in eICU-CRD.

| Prediction Time | Precision | Sensitivity | Specificity | F1 Score | ROC AUC | Accuracy |
| --- | --- | --- | --- | --- | --- | --- |
| 6 h Window | 0.623 | 0.774 | 0.974 | 0.690 | 0.971 | 0.964 |
| 12 h Window | 0.498 | 0.387 | 0.978 | 0.435 | 0.959 | 0.946 |
| 24 h Window | 0.270 | 0.411 | 0.930 | 0.326 | 0.904 | 0.900 |

